# A Bibliometric Analysis of Ophthalmology Resident Research Productivity in the United States

**DOI:** 10.1101/2022.02.06.22270335

**Authors:** Austin Huang, Sarah Kim, Harrison Zhu, Nihar Pathare, Xin Yee Ooi, R. Parker Kirby, Stephen P. Yoon, Zaina Al-Mohtaseb

**Affiliations:** Department of Ophthalmology, Baylor College of Medicine, Houston, TX

## Abstract

**Purpose:** To quantify scholarly activity of ophthalmology residents during residency and assess what factors may be associated with greater research productivity of these residents.

**Methods:** Ophthalmology residents who graduated in 2021 were identified from their respective program websites. Bibliometric data published by these residents between the beginning of their PGY-2 year (July 1st, 2018) until three months after graduation (September 30th, 2021) were captured through searches via PubMed, Scopus, and Google Scholar. The association of the following factors with greater research productivity numbers was analyzed: residency tier, medical school rank, sex, doctorate degree, type of medical degree, and international medical graduate status. Residency programs were sorted into five tiers by Doximity “reputation” and medical schools were stratified into *US News* Top 40 and non-Top 40 schools.

**Results:** We found 418 ophthalmology residents from 98 residency programs. These residents published a mean (±SD) number of 2.68±3.81 peer-reviewed publications, 2.39±3.40 ophthalmology-related publications, and 1.18±1.96 first-author publications each. The mean (±SD) h-index for this cohort was 0.79±1.17. Upon multivariate analysis, we discovered significant correlations between both residency tier and medical school rank and all bibliometric variables assessed. Pairwise comparisons revealed that residents from higher tier programs had greater research productivity numbers than those from lower tier programs.

**Conclusions:** We obtained bibliometric standards for ophthalmology residents on a national scale. Residents who graduated from higher-ranked residency programs and medical schools possessed higher *h*-indices and published more peer-reviewed publications, ophthalmology-related articles, and first author publications.

## INTRODUCTION

Scholarship is highlighted in all aspects of ophthalmology education. High impact research during medical school is associated with matching to a higher tier ophthalmology residency^1^, while ophthalmology fellowship directors list research as an important selection criterion for their programs.^2,3^ Furthermore, the Accreditation Council for Graduate Medical Education (ACGME) mandates participation in scholarship as a core tenet for residents at ophthalmology residency programs;^4^ however, the extent and impact of resident scholarly activity is not well understood.

Prior groups have used bibliometric analyses to examine research productivity among residency applicants in various fields, including ophthalmology.^1^ Other bibliometric studies have assessed resident scholarly output in various fields such as radiation oncology, neurosurgery, and general surgery.^5-7^ Here we sought to investigate the scope and significance of peer-reviewed research published by ophthalmology residents during residency in the United States. We conducted a bibliometric analysis to quantify the scholarly activity of ophthalmology residents and determine what factors may be associated with greater research productivity during residency.

## METHODS

We created a database of all ophthalmology residency programs in the United States, compiled from the ACGME website (www.acgme.org). Third-year (PGY-4) ophthalmology residents who graduated in 2021 were identified from the program websites of each institution. Residency programs were excluded from analysis if no ophthalmology residents graduated in 2021 or if the program websites lacked resident information.

We queried the PubMed database for all peer-reviewed publications published between the beginning of their ophthalmology residency (July 1st, 2018) and three months after graduation (September 30th, 2021) for each resident in the final cohort. The Scopus database and Google Scholar were cross-examined to ensure accuracy of publication list. Doximity and LinkedIn were used to confirm resident identities by checking undergraduate and medical schools, graduating year, and middle initials and names. Demographic information was compiled for each resident, including medical school institution, doctorate degree, type of medical degree, sex, and future plans if listed on the program website. Bibliometric variables collected for each resident included number of publications, number of ophthalmology-related publications, number of first-author publications, and *h*-index (manually measured), which is defined as the number of publications, *h*, that have been cited at least *h* times.^8^ Ophthalmology-related articles were divided by type of research into further categories: basic science, clinical research, case report, literature/systematic review, and other publications. Conference papers, abstracts, presentations, book chapters, and errata were excluded. IRB approval was not required for this retrospective database review.

We then conducted both univariate and multivariate analyses to determine potential factors that may be associated with greater research productivity among ophthalmology residents during residency. We analyzed the tier of residency programs that each resident graduated from and whether the resident attended a Top 40 medical school. We also looked at the following additional demographic factors: whether the resident was male or female, whether the resident had a doctorate degree (PhD) or not, whether the resident held an MD degree or a DO degree, and whether the resident was an international medical graduate (IMG) or not. The bibliometric variables that we assessed were number of publications, number of ophthalmology-related publications, number of first-author publications, and *h*-index. We divided all ACGME-accredited ophthalmology residency programs in the United States into five tiers (Tier 1: 1-20, Tier 2: 21-40, Tier 3: 41-60, Tier 4: 61-80, Tier 5: the remaining programs) by Doximity’s ranking by “reputation”. This ranking was created from the pooled results of surveys over the past three years that represent the opinion of ophthalmology board-certified Doximity members who ranked which programs provided the best clinical training.^9^ Medical schools were sorted into Top 40 and non-Top 40 categories in accordance with *US News*, which utilized total federal research activity as a part of its ranking methodology.^10^

Following a Kolmogorov-Smirnov test to determine if the variables followed a non-normal distribution, we performed a Kruskal-Wallis test comparing categorical predictors with quantitative responses on all bibliometric variables for association with residency tier, medical school rank, sex, doctorate degree, type of medical degree, and IMG status. Significant variables upon univariate analysis were used as inputs into a multivariate quasi-Poisson regression model, which corrected for overdispersed data.^11^ Dunn’s test with a Bonferroni correction was utilized to determine differences between residency program tiers for the variables that were found to be significant in the multivariate regression. Furthermore, we conducted multiple logistic regression exploring the relationship between quantitative predictors and a categorical response to determine whether there was a significant difference in resident research productivity and whether the resident decided to pursue a fellowship or practice comprehensive ophthalmology. Statistical analyses were performed in RStudio version 1.4.1106. Variables were found to be statistically significant at *P* < 0.05.

## RESULTS

A total of 124 ACGME-accredited ophthalmology programs were identified. Two programs were excluded for not graduating any residents in 2021, while a further 24 programs were excluded due to lack of resident information on their respective websites. In the remaining 98 residency programs, 418 ophthalmology residents who graduated in 2021 were identified. These residents published a mean (± SD) number of 2.68 ± 3.81 peer-reviewed publications each; the number of ophthalmology-related articles published was 2.39 ± 3.40, and the number of first author publications was 1.18 ± 1.96. The mean (± SD) *h*-index for this cohort of ophthalmology residents was 0.79 ± 1.17. Of the articles that were specific to the field of ophthalmology, 0.20 ± 0.61 publications were categorized as basic science, 1.13 ± 1.96 were related to clinical research, 0.75 ± 1.30 were case reports, and 0.32 ± 1.09 were literature or systematic reviews. These bibliometric variables were also sorted by our factors of interest: residency program tier, medical school rank (Top 40 or non-Top 40), sex, doctorate degree, type of medical degree, and IMG status (Table 1). Of the 418 residents identified, 238 of them had future plans listed on their respective residency program websites. The 176 residents pursuing a fellowship published a mean (± SD) number of 3.18 ± 4.44 publications, 2.84 ± 3.88 ophthalmology-related publications, and 1.41 ± 2.22 first author articles, with an *h*-index of 0.858 ± 1.29. The remaining 62 residents of this group planning to practice comprehensive ophthalmology had a mean (± SD) of 0.952 ± 1.21 peer-reviewed publications, 0.806 ± 1.02 articles in the field of ophthalmology, and 0.339 ± 0.745 first author publications published; their *h*-index was 0.371 ± 6.33.

**Table 1.** Means (and SD) of bibliometric variables sorted by selected demographic factors

|  | Number of residents | Publications | Ophthalmology publications | First author publications | H-index |
| --- | --- | --- | --- | --- | --- |
| All programs | 418 | 2.68 (3.81) | 2.39 (3.40) | 1.18 (1.96) | 0.79 (1.17) |
| Residency tier |  |  |  |  |  |
| Tier 1 | 116 | 4.10 (4.58) | 3.60 (4.46) | 1.91 (2.65) | 1.22 (1.31) |
| Tier 2 | 90 | 3.22 (3.98) | 3.02 (3.68) | 1.30 (1.85) | 0.89 (1.18) |
| Tier 3 | 89 | 2.01 (3.83) | 1.66 (2.40) | 0.94 (1.66) | 0.56 (0.81) |
| Tier 4 | 71 | 1.61 (1.76) | 1.44 (1.70) | 0.59 (0.94) | 0.68 (1.40) |
| Tier 5 | 52 | 1.17 (2.12) | 1.14 (2.13) | 0.54 (1.18) | 0.25 (0.52) |
| Top 40 | 138 | 3.88 (4.98) | 3.39 (4.40) | 1.72 (2.54) | 1.14 (1.21) |
| Non-Top 40 | 280 | 2.09 (2.92) | 1.90 (2.66) | 0.91 (1.53) | 0.62 (1.12) |
| Male | 265 | 2.37 (3.45) | 2.18 (3.36) | 1.07 (2.00) | 0.73 (0.97) |
| Female | 153 | 3.20 (4.33) | 2.76 (3.47) | 1.37 (1.87) | 0.92 (1.46) |
| PhD | 12 | 4.33 (2.93) | 3.25 (3.08) | 1.58 (1.24) | 1.50 (1.17) |
| No PhD | 406 | 2.63 (3.83) | 2.37 (3.42) | 1.17 (1.98) | 0.77 (1.17) |
| MD | 405 | 2.73 (3.86) | 2.44 (3.45) | 1.20 (1.98) | 0.82 (1.18) |
| DO | 13 | 1.00 (1.00) | 1.00 (1.00) | 0.62 (0.77) | 0.08 (0.28) |
| IMG | 19 | 5.90 (5.05) | 4.84 (4.62) | 2.16 (2.41) | 1.84 (1.50) |
| Non-IMG | 399 | 2.52 (3.68) | 2.27 (3.30) | 1.13 (1.93) | 0.74 (1.13) |

The Kolmogorov-Smirnov test showed that all bibliometric variables were skewed to the right and not normally distributed, with Figure 1A displaying the distribution of the number of first author publications published by each resident and Figure 1B exhibiting the distribution of index per resident. Upon univariate analysis, the tier of residency program the resident graduated from, whether the resident attended a Top 40 or non-Top 40 medical school, and whether the resident was an international medical graduate (IMG) or not were found to be significantly associated with all bibliometric variables analyzed: number of publications, number of ophthalmology-related publications, number of first author publications, and *h*-index. Furthermore, residents with a doctorate degree possessed higher *h*-indices (*P* = 0.012) and published significantly more publications (*P* = 0.007) than those who did not have a PhD, while female residents published significantly more articles as first author than their male counterparts (*P* = 0.046) and MD residents obtained significantly higher *h*-indices than those with DO degrees (*P* = 0.004). Multivariate analysis revealed only residents who graduated from higher tier residencies, attended Top 40 medical schools, or were international medical graduates had significantly higher research productivity numbers for all bibliometric variables of interest (Table 2). Dunn’s test with a Bonferroni correction (adjusted *P* < 0.05) was conducted to show that residents who graduated from a Tier 1 residency program had significantly higher numbers of publications, ophthalmology-related publications, first author publications (Fig. 2A), and *h*-index (Fig. 2B) when compared to graduates of Tier 3, 4, 5 programs, as well as significantly higher numbers for these bibliometric variables for graduates of Tier 2 as opposed to those of Tier 5 residencies. The adjusted *P* values for these findings are summarized in Table 3. Additionally, a multiple logistic regression model found that no bibliometric variables were statistically significant predictors for choosing to pursue a fellowship as opposed to practicing comprehensive ophthalmology.

**Table 2.** Univariate and multivariate effects between bibliometric variables and selected demographic factors

|  | Publications | Ophthalmology publications | First author publications | H-index |
| --- | --- | --- | --- | --- |
| P-value (univariate) |  |  |  |  |
| Residency tier | <0.001* | <0.001* | <0.001* | <0.001* |
| Medical school rank | <0.001* | <0.001* | <0.001* | <0.001* |
| Sex | 0.092 | 0.104 | 0.046* | 0.667 |
| Doctorate degree | 0.012* | 0.259 | 0.052 | 0.007* |
| Medical degree type | 0.097 | 0.182 | 0.494 | 0.004* |
| IMG status | <0.001* | 0.006* | 0.013* | <0.001* |
| P-value (multivariate) |  |  |  |  |
| Residency tier | <0.001* | <0.001* | <0.001* | <0.001* |
| Medical school rank | <0.001* | 0.004* | 0.011* | 0.004* |
| Sex |  |  | 0.516 |  |
| Doctorate degree | 0.878 |  |  | 0.483 |
| Medical degree type |  |  |  | 0.199 |
| IMG status | <0.001* | <0.001* | <0.001* | <0.001* |

\*Statistically significant, $p < 0.05$

**Table 3.** Multiple pairwise comparisons between residency program tier and bibliometric variables

| Residency tier | Tier 1 | Tier 2 | Tier 3 | Tier 4 | Tier 1 | Tier 2 | Tier 3 | Tier 4 |
| --- | --- | --- | --- | --- | --- | --- | --- | --- |
| Number of publications |  |  |  |  | Number of ophthalmology publications |  |  |  |
| Tier 2 | 0.524 |  |  |  | 1.00 |  |  |  |
| Tier 3 | <0.001* | 0.126 |  |  | <0.001* | 0.057 |  |  |
| Tier 4 | <0.001* | 0.184 | 1.00 |  | 0.001* | 0.061 | 1.00 |  |
| Tier 5 | <0.001* | <0.001* | 0.782 | 0.932 | <0.001* | <0.001* | 1.00 | 1.00 |
| Number of first author publications |  |  |  |  | H-index |  |  |  |
| Tier 2 | 0.770 |  |  |  | 0.276 |  |  |  |
| Tier 3 | 0.002* | 0.706 |  |  | 0.001* | 1.00 |  |  |
| Tier 4 | <0.001* | 0.085 | 1.00 |  | 0.008* | 1.00 | 1.00 |  |
| Tier 5 | <0.001* | 0.017* | 1.00 | 1.00 | <0.001* | 0.003* | 0.236 | 0.188 |

\*Statistically significant, Bonferroni-adjusted $p < 0.05$

**Figure 1.**
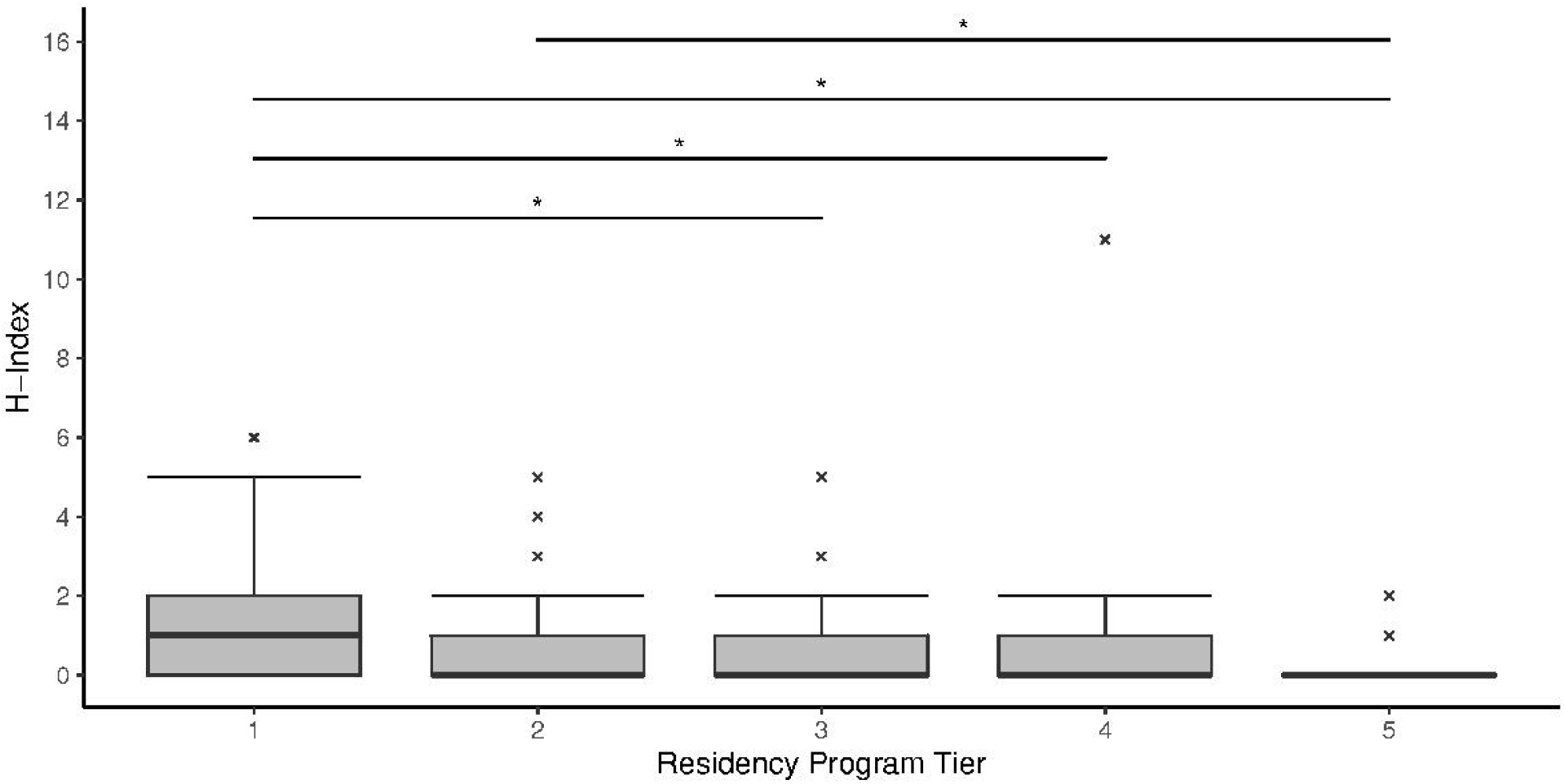
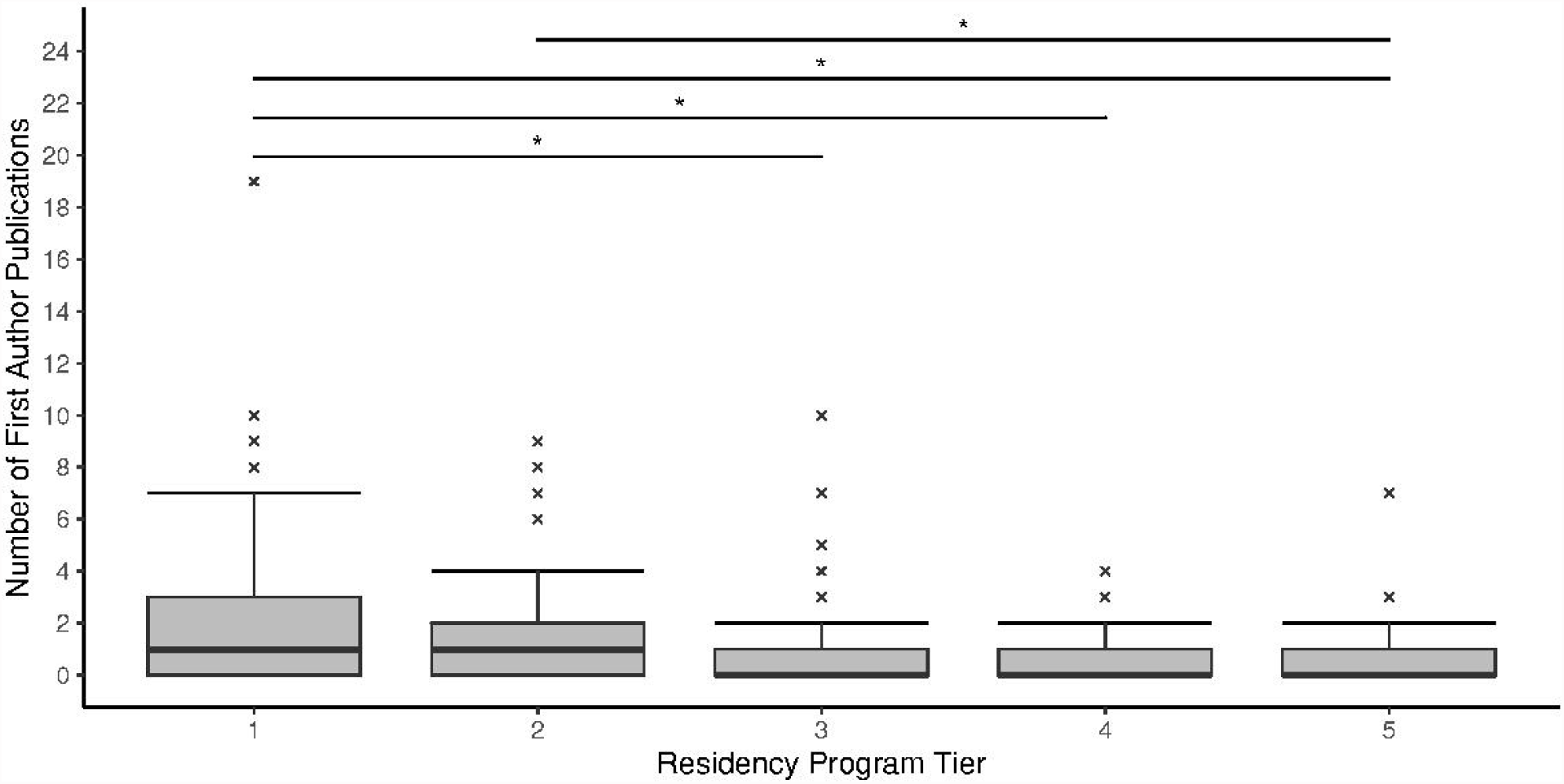
Right-skewed distribution of **(A)** number of first author publications for each resident and **(B)** *h*-index for each resident. The number of residents per bar is shown.

**Figure 2.**
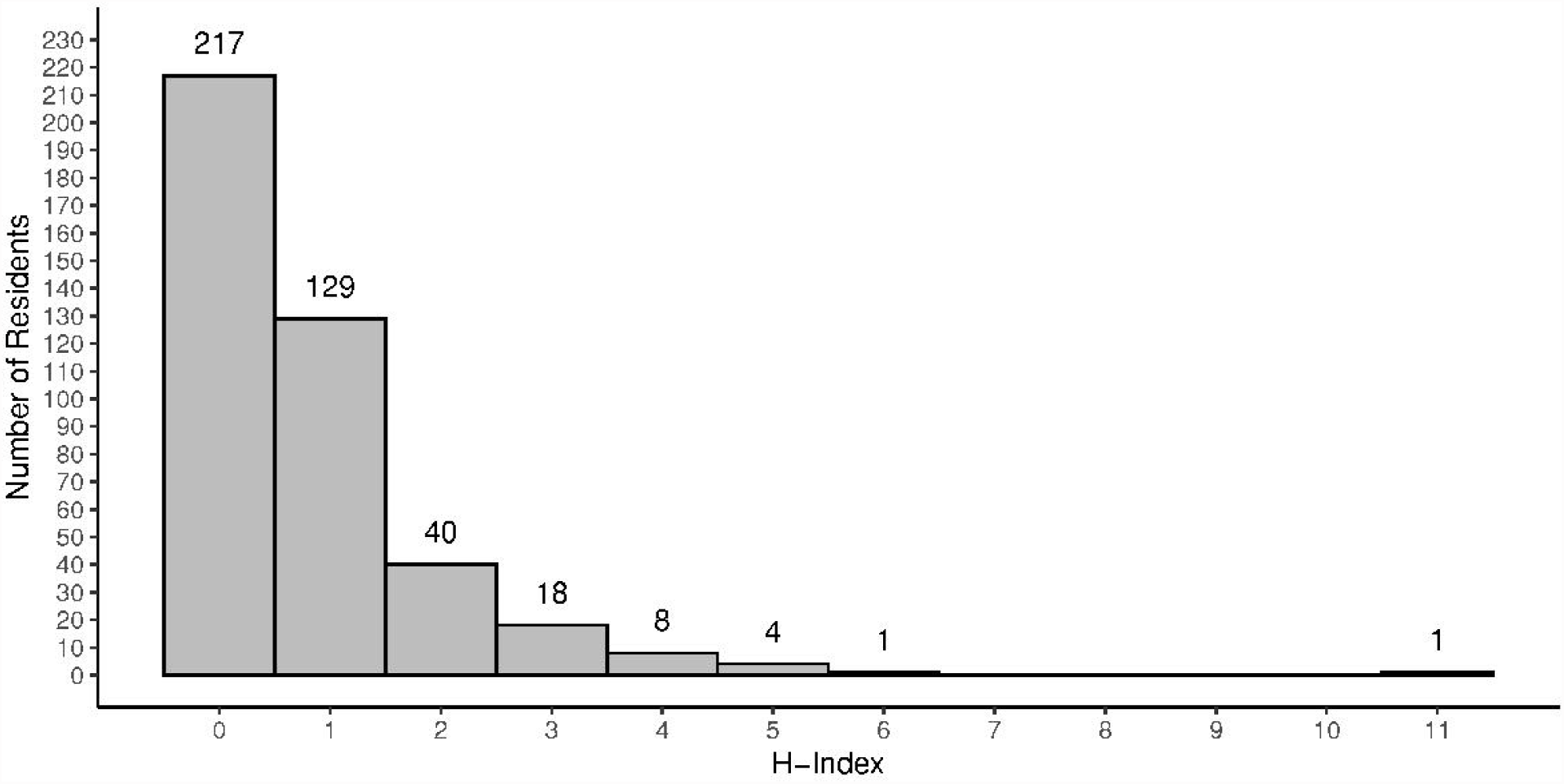
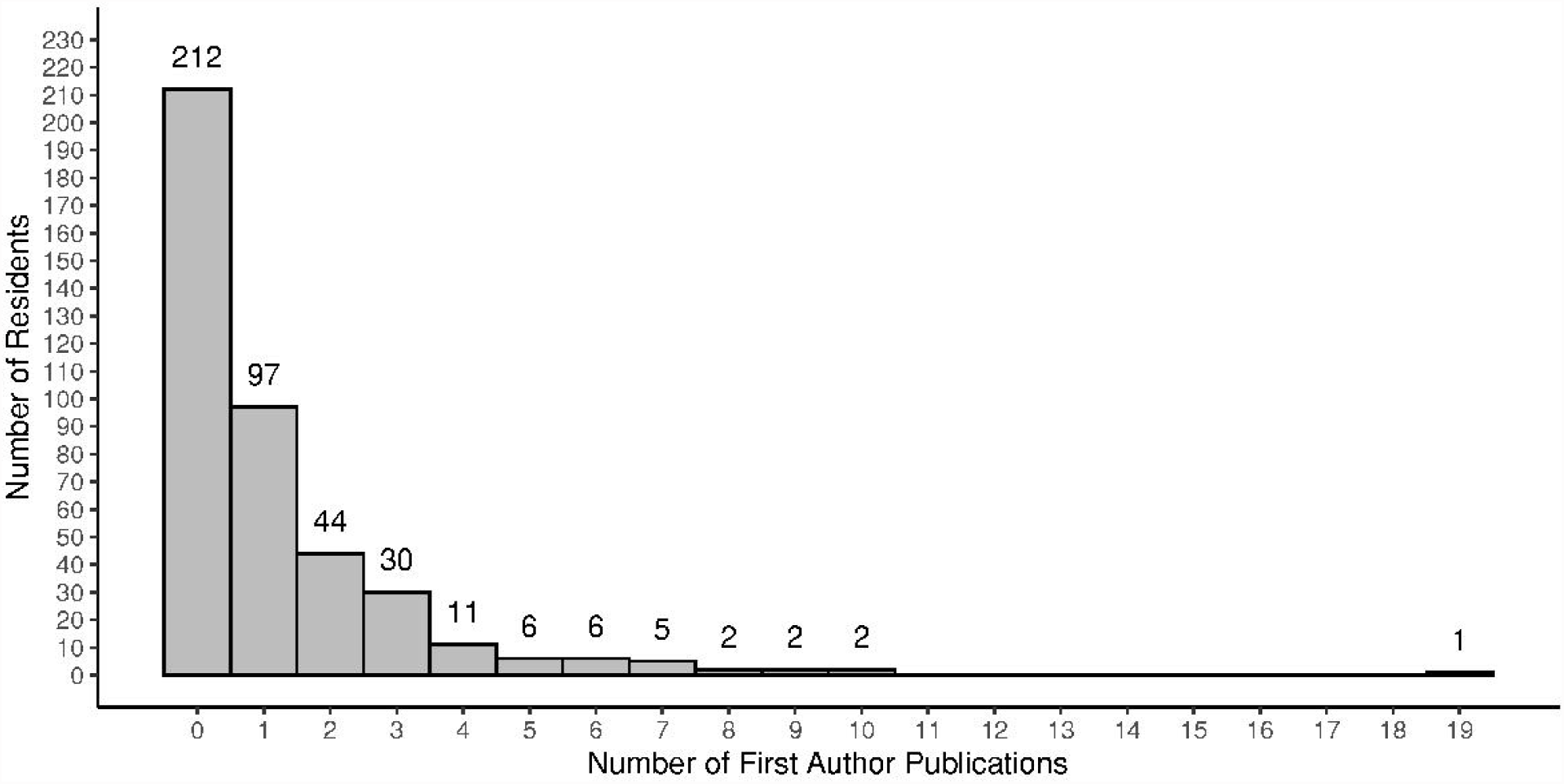
Box-plots of **(A)** the number of first author publications per resident and **(B)** *h*-index per resident compared with residency program tier from which the resident graduated. Dunn’s test with a Bonferroni correction revealed that Tier 1 residents possessed higher *h*-indices and produced a greater number of publications, ophthalmology-related publications, and first author publications in comparison to those of Tier 3, 4, 5 programs. The same significance was found between residents from Tier 2 and Tier 5 residencies. Adjusted *P* < 0.05 and outliers labeled by (x).

## DISCUSSION

Upon multivariate analysis, we found a significant association between both the program tier from which a resident graduated and the rank of medical school they attended and their research productivity during residency. While not all the pairwise comparisons between the different program tiers yielded significant results, residents who graduated from higher-ranked residencies obtained higher *h*-indices and produced more publications, ophthalmology-related publications, and first author publications than their peers who graduated from lower-ranked programs did. Likewise, ophthalmology residents who attended Top 40 medical schools produced significantly higher numbers during residency for all bibliometric variables assessed than those who graduated from non-Top 40 medical schools. Similarly, we found that residents who graduated from international medical schools obtained higher research productivity numbers than those who did not. However, upon multivariate analysis, there were no significant differences among research productivity numbers between male and female residents, whether they had a doctorate degree or not, and whether they held an MD or a DO degree. Furthermore, none of these bibliometric variables were significant predictors for whether residents chose to pursue fellowship or practice comprehensive ophthalmology after graduation from residency following multivariate analysis.

Higher-ranked ophthalmology residency programs may have more resources and opportunities for resident research, which contributes to the result that residents who graduated from residencies in higher tiers produced higher research productivity numbers than those who graduated from residencies in lower tiers did. These programs may also emphasize academic research as a larger part in the training of ophthalmology residents. Likewise, we found that residents who attended Top 40 medical schools were more productive during residency than those who attended non-Top 40 schools. This metric remained statistically significant upon multivariate analysis; thus, it may be possible that an early exposure to research opportunities is correlated with greater research productivity in the future, since the Top 40 schools possess more resources, such as federal funding, than their non-Top 40 counterparts.

An interesting aspect of our results is that we found no significant correlation between sex and research productivity for any of the bibliometric variables assessed. In the bibliometric analyses of both radiation oncology and neurosurgery residents, male residents seemed to produce higher numbers of research productivity metrics when compared to their female counterparts.^5,6^ However, two recent studies on general surgery and plastic surgery residents revealed that there was no significant difference between male and female residents.^7,12^ It is possible that there is a potential change underway in terms of increasing female representation in academic publications, which counteracts a long-standing trend of male dominance in academia that has been noted across various specialties, including ophthalmology.^13^

We also found no significant associations between whether the resident had a doctorate degree or whether they held an MD or a DO degree. We hesitate to draw conclusions from a small sample size, as neither are well-represented among ophthalmology residents: 12 residents had a PhD and 13 residents held a DO degree. However, the difficulty of matching into a competitive specialty as an osteopathic medical graduate as opposed to an allopathic one is well-known (18% of DO applicants successfully matched into ophthalmology residency programs as opposed to 75% of MD applicants).^14^

On the other hand, while only 19 residents were IMG’s, we found that residents who graduated from international medical schools have significantly greater research productivity during residency compared to their counterparts who did not. International medical graduates often face a non-traditional path to obtaining a residency spot in the United States, including graduating from prior residencies abroad and participating in pre-residency research fellowship programs; these prior research experiences may be contributing factors to why IMG research productivity during residency is greater than that of graduates from medical schools in the United States.^15^

We also considered the possibility that research productivity affected whether the resident pursued a fellowship as opposed to practicing comprehensive ophthalmology after graduation; however, no significant outcomes were obtained. Only 238 out of a total 418 residents had their future plans listed on their respective residency program websites, and this result may not be reflective of the graduating class of 2021 as a whole.

To the best of our knowledge, this is the first bibliometric study to characterize and analyze the research productivity of ophthalmology residents during residency. Other groups have utilized bibliometric analyses to examine research productivity in other residencies, such as radiation oncology, neurological surgery, and general surgery.^5-7^ Additional studies have used bibliometric data to assess correlations between medical school research productivity and matching into a specific specialty’s residency programs, including ophthalmology.^1,16-19^ We modified the protocols employed in these studies for the collection of bibliometric data, as well as the methods utilized for statistical analysis, to ensure the reliability and validity of our conclusions. By cross-referencing the Scopus database and Google Scholar with PubMed, we attempted to capture all peer-reviewed, indexed publications during the given time frame: the beginning of their ophthalmology residency (July 1st, 2018) until three months after graduation (September 30th, 2021). Furthermore, we used both Doximity and LinkedIn to confirm resident identities to ensure accuracy of the bibliometric data collected.

There were several limitations to note in this study. First, 24 programs were excluded for lacking resident information on their respective websites. However, 418 graduated residents were identified from the remaining 98 residency programs, which is 90.5% of the class that matched into ophthalmology residency spots in January 2017, giving us confidence that our data reflects the research productivity of the vast majority of the graduating class of 2021.^20^ Second, the methodology by which Doximity calculates its residency rankings by “reputation” is dependent on the responses of survey-eligible ophthalmologists.^9^ Even though there is an inherent bias to these rankings, they are often used to separate residency programs into tiers for analysis.^1,16-18^ Similarly, *US News* determines research rankings for medical schools based in part on how much federal funding each school receives.^10^ Despite its flaws, other bibliometric studies have also used the Top 40 stratification for medical schools as a variable.^16^ Third, although we found no significant differences in research productivity for residents who are pursuing a fellowship as opposed to comprehensive ophthalmology, in addition to assessing demographic factors such as sex, there may be further confounding factors that affect scholarly activity among residents during residency. Finally, we only captured peer-reviewed, indexed publications published by residents between July 1st, 2018 and September 30th, 2021, as described in prior studies.^5^ Since the publishing process can often be long, there is a high likelihood that there are projects that residents worked on during residency but were not published until after our end date. Although we used three different databases to collect bibliometric data, some publications may not be indexed or peer-reviewed. Likewise, research productivity is not only defined by peer-reviewed publications; rather, we excluded conference papers, abstracts, presentations, book chapters, and errata, which are difficult to objectively capture but may contribute to valuable research experience. Furthermore, while *h*-index is a well-known tool to quantify the impact of scholarly output, it does not consider the importance of author position.^8^ We included number of first author publications as an individual bibliometric variable to account for this limitation. Even though we determined that there is no significant difference between male and female residents in terms of research productivity, it may be challenging to conduct a thorough bibliometric search if a resident changed the surname they use for authorship. Despite our best efforts to confirm author identities, there is a possibility that there are peer-reviewed publications published by residents that are unaccounted for in this study.

In conclusion, we characterized scholarly activity of United States ophthalmology residents during residency in an objective manner and assessed factors that are associated with greater research productivity during residency with statistical robustness. We obtained bibliometric standards for residents in ophthalmology programs on a national scale and subdivided our cohort into several important demographic distinctions. Notably, we discovered that residents who graduated from higher-ranked residency programs and medical schools possessed higher *h*-indices and published more peer-reviewed publications, ophthalmology-related articles, and first author publications. With the results of this bibliometric study, we hope to better inform residents, medical students, residency and fellowship program directors, and potential future employers about current trends in scholarly activity.

## Data Availability

The data used to support our findings were obtained from publicly available websites as detailed in our methods section; no new data was generated through this study.

## Other Acknowledgements

None

